# eREACH: A randomized trial of digital alternatives to genetic counseling for metastatic cancer

**DOI:** 10.1101/2025.10.01.25337108

**Authors:** Kimberley T. Lee, Brian Egleston, Briana McLeod, Sarah Brown, Sarah Howe, Dominique Fetzer, Susan M. Domchek, Lauren Gutstein, Cara Cacioppo, Dana Clark, Janice Christiansen, Jessica Ebrahimzadeh, Demetrios Ofidis, Hannah Griffin, Rajia Mim, Santina Hernandez, Linda Fleisher, Kelsey Karpink, Enida Selmani, Aysha Tahsin, Evelyn Mastaglio, Lynne Wagner, Michelle Weinberg, Kuang Yi-Wen, Elisabeth Wood, Angela R. Bradbury

**Affiliations:** University of Pennsylvania, Division of Hematology-Oncology; University of Pennsylvania, Abramson Cancer Center; Fox Chase Cancer Center, Department of Biostatistics and Bioinformatics; Fox Chase Cancer Center, Cancer Prevention and Control; Wake Forest School of Medicine, Department of Health Policy and Management; Thomas Jefferson University Hospital, Department of Medical Oncology; University of Pennsylvania, Department of Medical Ethics and Health Policy

## Abstract

With FDA approval of targeted therapies in patients with germline BRCA1/2-related advanced cancers there is a need to evaluate efficient and effective delivery models for germline cancer genetic testing. We sought to evaluate the effectiveness of replacing traditional pretest and posttest counseling with a genetic counselor (GC) with a digital intervention in patients with metastatic cancers. eREACH is a 4-arm randomized non-inferiority trial with a 2×2 design conducted from 2020-2024 and participants were followed for 6 months. Participants were recruited from the community and academic and community medical sites across the United States. A referred sample of adults with advanced or metastatic breast, prostate, ovarian or pancreatic cancer. The digital pretest intervention consists of 8 modules including purpose of genetic testing and implications of results. The digital post-test intervention consists of 4 modules including test results and explanation of results. The primary endpoint was non-inferiority in change in knowledge and anxiety from baseline to post-disclosure. Secondary analyses evaluated patient-reported outcomes (PROs) such as depression and distress, and moderators of PROs such as socioeconomic status. 229 participants were recruited from 14 states and 37% were male, 17% were non-white, 43% had less than a college education and 21% were from community sites or national recruitment. We met non-inferiority for all short-term PROs for both one visit arms (GC/Digital, Digital/GC). We met non-inferiority for all short-term PROs in the fully digital arm except knowledge which was inconclusive, although differences were small. Uptake of visit 1 was lower in the digital arms, although uptake of testing after visit 1 did not differ between arms. Patients living in poorer areas had greater reductions in anxiety in the visit 1 GC arms and rural patients had greater increase in knowledge in digital arms. Men had greater decreases in anxiety with digital disclosure, while women had greater reductions with GC disclosure. The eREACH patient-centered digital delivery intervention with one GC visit is an evidence-based alternative to two visits with a GC for patients with metastatic cancer. The fully digital model may be acceptable for some patients.

## INTRODUCTION

Germline cancer genetic testing has important therapeutic implications for patients with advanced breast, ovarian, pancreatic, and prostate cancers. The Food and Drug Administration (FDA) has approved poly-ADP ribose polymerase (PARP) inhibitors as treatment for patients with these cancers in the setting of a germline *BRCA 1/2* pathogenic or likely pathogenic variant (PV). Germline *BRCA* PV are also associated with increased sensitivity to platinum chemotherapeutic agents. Patients with prostate cancer and any deleterious germline homologous recombination repair mutation may also qualify for treatment with PARP inhibitors. Pembrolizumab is approved for patients with microsatellite instability high (MSI-H) cancers and more than 90% of Lynch syndrome tumors are MSI-H. Yet, many at-risk patients do not have access to genetic services, leaving many genetic carriers unidentified.^1–5^ Access to genetic counselors (GC) is limited in many areas in the US, and the traditional delivery model of pre- and post-test counseling with a genetic professional will not support the rising indications for germline genetic testing. Thus, there is a pressing need to evaluate alternative delivery models to increase access to and efficiency of germline testing for therapeutic indications.

To address this need, we developed eREACH, a 2×2 non-inferiority study randomizing patients to a patient-centered multimodal digital intervention versus traditional pre-test (visit 1: education and consent) and/or post-test (visit 2: disclosure) counseling delivered by a GC. The primary objective was to assess whether the digital intervention would result in equal or improved cognitive and affective outcomes (knowledge and anxiety) from baseline to post-disclosure assessment. Secondary outcomes include cognitive, affective, and behavioral outcomes from baseline to post-disclosure and at 6 months.

## MATERIALS AND METHODS

### Participants

Participants were English speaking adults diagnosed with metastatic breast or prostate cancer, and metastatic or advanced ovarian or pancreatic cancer. Participants were recruited from Penn Medicine oncology clinics, regional community oncology clinics affiliated with the Penn Telegenetics program, and social media ads through Penn and Cancer Support Community, a cancer advocacy organization. Patients with prior germline genetic testing, uncorrected or uncompensated speech defects, uncontrolled psychiatric/mental condition or cognitive deficits rendering the individual unable to understand study goals or tasks were excluded. Remote genetic counseling services in this study were provided through 8 GCs in the Penn Telegenetics Program and Penn Cancer Risk Evaluation Program. The study protocol was approved by the University of Pennsylvania’s institutional review board.

### Digital Interventions for Pre-test education and disclosure of results

As previously described, the eREACH digital interventions are theoretically informed, user-tested, interactive patient-centered digital intervention for germline genetic testing in advanced or metastatic cancers. Intervention details are described using the GeM-CheckD checklist^6^ (**Supplemental Table 1**) and specific modules and content have been previously described.^7^ Briefly, the digital intervention includes Tier 1, indispensable information presented to all users, and optional Tier 2 content (more in-depth information, examples and/or videos). The pre-test intervention included 8 modules and participants were asked to record their testing decision. The digital result disclosure (visit 2) intervention includes 4 modules as previously described.

### Randomization and Procedures

After informed consent and the baseline survey (T0), participants were randomized to one of the four arms, stratified by gender and cancer type and using a permuted block design.

In the intervention arms (Arms B-D) the traditional standard-of-care pre-test (visit 1) and/or post-test (visit 2) counseling delivered by a GC was replaced with the digital intervention to give Arms A (GC/GC), B (GC/digital), C (digital/GC) and D (digital/digital). Participants assigned to a digital visit could request a visit with a GC if preferred. All visits with a GC were conducted by phone or videoconference. Full study procedures have been previously described.^7^

All genetic testing was sent to a commercial laboratory and covered by insurance. Participants could choose a targeted panel limited to genes which could immediately impact treatment for metastatic cancer (e.g. *BRCA1/2* and Lynch Syndrome), a larger gene panel which included additional genes related to their cancer type which could have an impact on treatment in the future or additional impact for relatives, or a customized panel based on other health concerns or a desire to exclude certain genes (see **Supplemental Table 2**).

### Primary Outcomes

Theoretically informed outcomes to evaluate cognitive, affective and behavioral outcomes of digital alternatives as compared to meeting with a GC were collected at baseline (T0), after Visit 1 (T1) and Visit 2 (T2) and at 6 months (T3). Participants received a $10 gift card for each survey completed.

The primary outcomes included:

1. *Knowledge of genetic disease* evaluated with the 16-item The KnowGene Scale (Cronbach α = 0.83).^8^
2. Anxiety assessed by the 4-item Patient Reported Outcomes Measurement Information System (PROMIS) Anxiety measure (Cronbach α = 0.79).^9^

### Secondary Outcomes

Secondary outcomes have been previously described^7^ and included: 1) Uptake of counseling and testing, 2) Communication of results, 3) Treatment change as a result of genetic test results (determined via chart review), and PROs 4) Depression (4-item PROMIS Depression, alpha=0.86-0.96),^9^ 5) Disease-specific distress (Impact of Events Scale, alpha = 0.82-0.90),^10–13^ 6)Satisfaction with genetic services, 7) Responses to testing (Multi-dimensional Impact of Cancer Risk Assessment Questionnaire, Cronbach α = 0.88),^14^ and 8) Decisional regret (5-item Decision Regret Scale, Cronbach α = 0.81-0.92).^15,16^ GCs recorded time spent on patient care activities for a subset of participants to estimate provider time by arm.

Moderators were collected at baseline and included race/ethnicity, education, marital status, gender and age. The Social Vulnerability Index (SVI) is a place-based index designed to quantify communities experiencing social vulnerability based on socioeconomic status, household characteristics, racial and ethnic minority status, and housing type and transportation and was identified using participant zip code.^17^ Higher scores indicate greater social vulnerability. **Health literacy was** assessed with the Brief Health Literacy Screen (Cronbach α = 0.81-0.79).^18^ Family history of cancer was defined as cancers of the breast, ovary, pancreas, or colorectum in a first or second-degree relative.

### Statistical Analysis

The primary outcomes included changes in knowledge and anxiety from the baseline to the post-disclosure period (T0-T2). Secondary outcomes included uptake of testing, depression, cancer specific distress, uncertainty, change in treatment plan, communication of results and provider time. Our primary non-inferiority test was based on an ANOVA test. We declared noninferiority under two conditions: 1) if the joint p-value>0.15 and all three standardized effects comparing arms B-D to arm A did not exceed 0.25 standard deviations in the inferior direction, or 2) if the joint p-value≤0.15 and the bound of the one-sided 85% confidence interval did not cross 0.35 standard deviation units in the inferior direction. Our design had >80% power and <1% Type I error under hypothesized scenarios. We standardized based on pooled baseline data. We additionally compared arms using T-tests and chi-squared tests. We used linear and logistic regressions to assess moderators of intervention effects.

## RESULTS

### Study Participants

Enrollment, randomization, and survey completion are shown in **Figure 1**. Of 305 eligible participants, 256 individuals consented to the study (83.9% of eligible) and 229 (89.4% of eligible consented) completed the baseline survey and were randomized. There were no statistically significant baseline differences between randomized groups (**Table 1**). The mean age of the participants was 67 years, 83.4% were white, 10% were Black, 3.1% were Hispanic/Latinx, and 42.9% had less than a college degree. Participants were recruited from 14 states across the United States, 38.6% were from rural areas, 21% were referred from a community site and 8.7% were recruited from social media or otherwise self-referred.

**Table 1:**
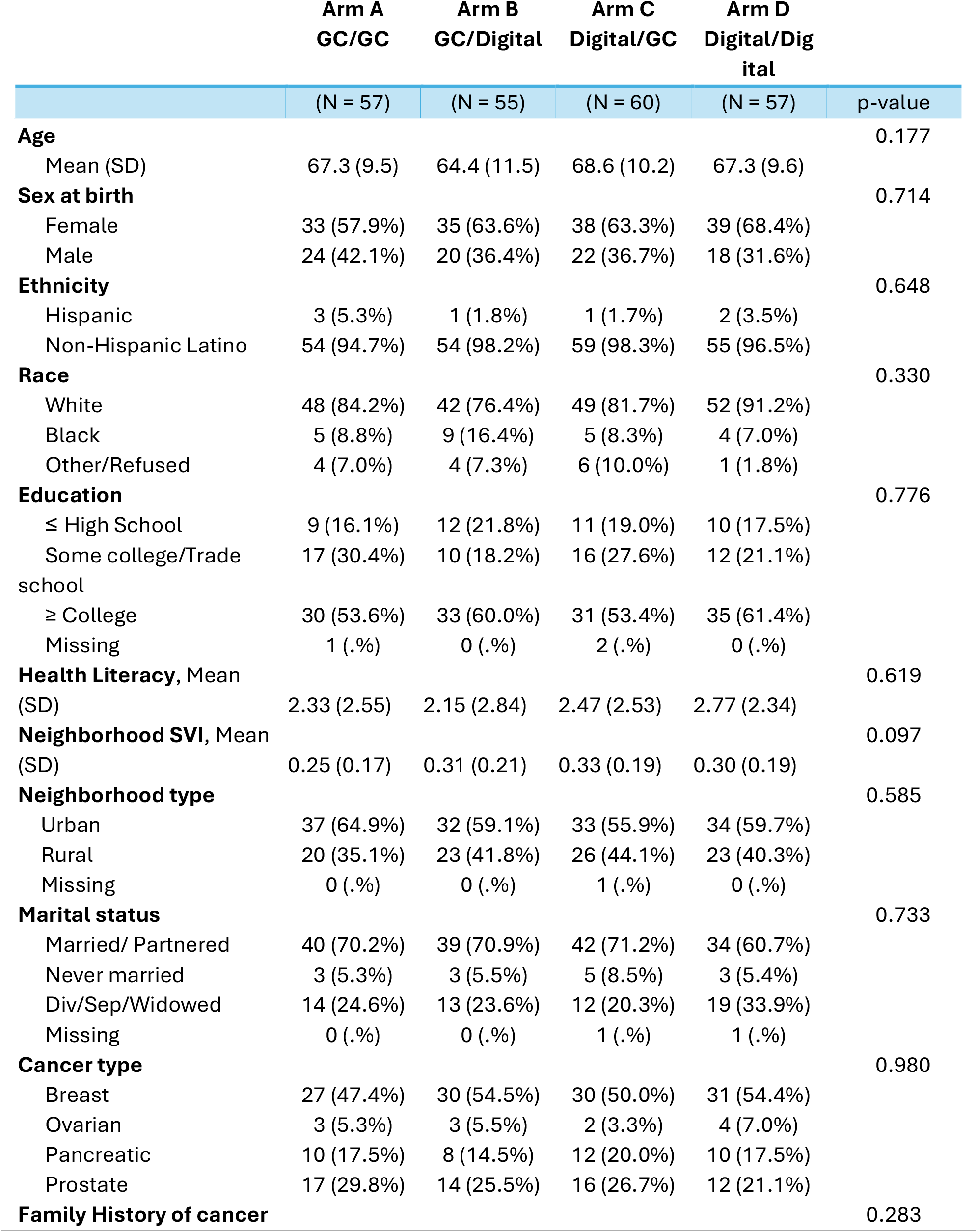
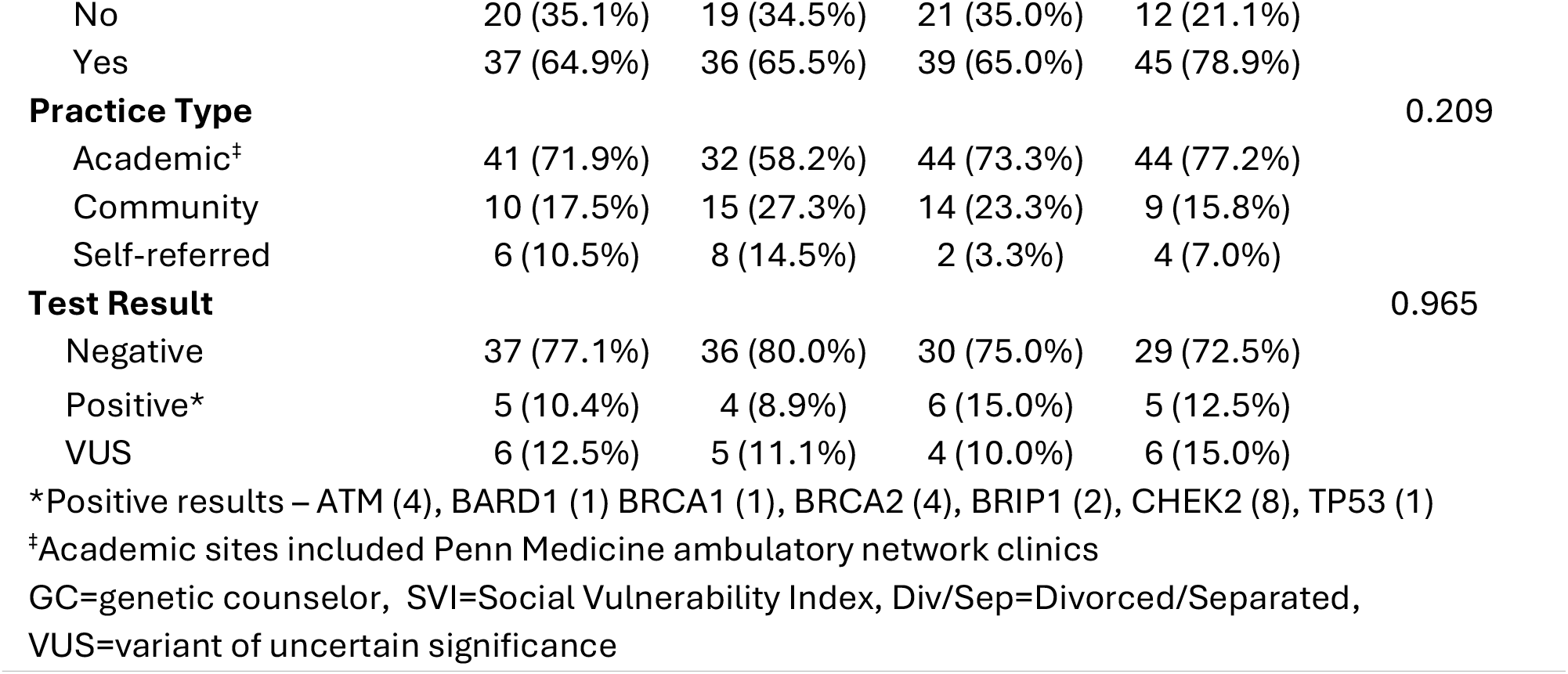
Participant Characteristics at Baseline by Study Arm.

**Figure 1.**
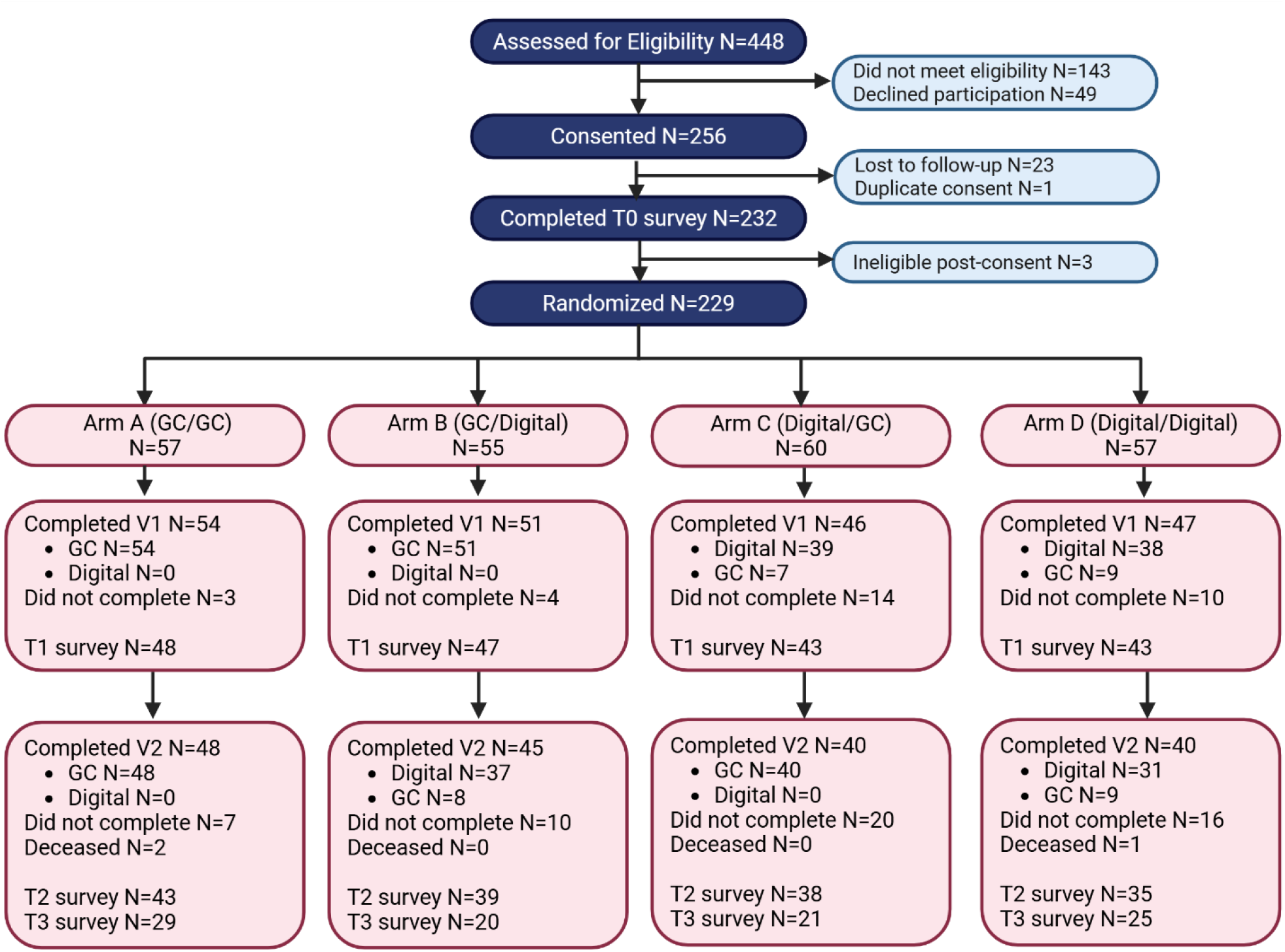
Consolidated Standards of Reporting Trials diagram. Survey completion rates: T1 181/229 (79%), T2 155/229 (68%), T3 95/229 (41 %) Participants spent an average of 31.8 minutes (range 3.7‐128.2 minutes) completing digital visit 1 with an average number of total logins of 1.7 (range 1‐6). The average time spent completing digital visit 2 was 6.2 minutes (range 0.2‐40.2 minutes). The average number of total logins for visit 2 was 1.7 (range 1‐9).

Most participants (63.3%) chose a larger panel, 12.2% chose a targeted panel, and 10% chose a custom panel. Neither demographic characteristics nor cancer type were associated with testing choices. Most participants (76.3%) had a negative result, 20 (11.6%) had a positive result and 21 (12.1%) had a VUS.

### Non-inferiority analyses and uptake of testing

In the primary intention-to-treat (ITT) analyses, we met the non-inferiority thresholds for anxiety (T0-T2) and all secondary short-term PROs (T0-T1, T0-T2) in all arms (**Figure 2**). For the primary outcome of knowledge (T0-T2), we met the non-inferiority thresholds in arms B and C, but non-inferiority was inconclusive for arm D. As shown in **Table 2**, increases in knowledge (T0-T2) were greater in Arms A-C as compared to Arm D, although differences were small and not statistically significant. In the ITT analyses of our primary 6-month outcomes (T0-T3), we met non-inferiority thresholds for anxiety for all arms and for knowledge in Arms B and D, but non-inferiority for knowledge was inconclusive for Arm C (**Supplemental Figure 1**). We met the non-inferiority thresholds for all for all 6-month secondary PROs except disease specific distress in Arm D.

**Table 2:** Mean differences (standard deviation) by arm for the primary and secondary outcomes after visit 1 (T1), visit 2 (T2), and at 6 months (T3).

| Outcomes, mean (SD) | Arm A (n=57) | Arm B (n=55) | Arm C (n=60) | Arm D (n=57) | p<sup>a</sup> |
| --- | --- | --- | --- | --- | --- |
| Anxiety (T1-T0) | -1.10 (2.17) | -1.83 (2.51) | -1.49 (2.89) | -0.67 (2.15) | 0.138 |
| Anxiety (T2-T0) | -1.93 (2.32) | -2.36 (2.43) | -2.32 (3.09) | -1.83 (2.83) | 0.732 |
| Anxiety (T3-T0) | -0.59 (2.77) | -0.85 (2.66) | -0.62 (2.64) | -1.00 (2.92) | 0.944 |
| Knowledge (T1-T0) | 2.92 (3.05) | 2.71 (4.38) | 2.56 (2.93) | 2.71 (3.73) | 0.971 |
| Knowledge (T2-T0) | 2.47 (3.35) | 2.62 (4.15) | 2.03 (3.29) | 1.23 (3.49) | 0.349 |
| Knowledge (T3-T0) | 1.96 (3.51) | 2.13 (3.21) | 0.88 (3.43) | 1.89 (3.33) | 0.619 |
| Depression (T1-T0) | -0.42 (1.83) | -0.55 (1.56) | -0.58 (1.69) | -0.35 (2.07) | 0.918 |
| Depression (T2-T0) | -0.91 (1.99) | -0.74 (1.89) | -0.92 (1.82) | -1.26 (2.73) | 0.769 |
| Depression (T3-T0) | -0.21 (1.93) | 0.10 (2.22) | 0.19 (2.14) | 0.36 (2.46) | 0.811 |
| Disease specific distress (T1-T0) | -1.80 (7.18) | -1.94 (7.20) | -1.41 (6.85) | -1.41 (5.67) | 0.975 |
| Disease specific distress (T2-T0) | -2.33 (6.42) | -2.30 (9.31) | -2.10 (7.79) | -1.44 (7.62) | 0.958 |
| Disease specific distress (T3-T0) | -1.60 (6.82) | -2.45 (7.18) | -0.79 (4.93) | 1.29 (7.74) | 0.274 |
| Satisfaction with services (T1) | 43.33 (4.20) | 42.36 (5.03) | 43.01 (4.47) | 42.66 (3.80) | 0.788 |
| Satisfaction with services (T2) | 42.75 (4.25) | 43.15 (4.84) | 42.67 (5.26) | 43.99 (3.86) | 0.604 |
| Decisional regret(T2) | 8.37 (12.33) | 11.58 (14.62) | 6.71 (10.73) | 10.43 (13.69) | 0.364 |
| Responses to testing (MICRA, T2) | 11.90 (9.37) | 10.13 (9.39) | 8.76 (9.20) | 13.99 (11.99) | 0.135 |
| Responses to testing (MICRA, T3) | 13.29 (8.79) | 12.88 (7.59) | 12.09 (7.54) | 14.94 (10.37) | 0.729 |
| Uptake of Visit 1, n, (%) | 54 (94.7%) | 51 (92.7%) | 46 (76.7%) | 47 (82.5) | 0.012 |
| Uptake of testing n (%) | 48 (84.2%) | 45 (81.8%) | 40 (66.7%) | 40 (70.2%) | 0.074 |
| Uptake of Visit 2, n (%) | 48 (84.2%) | 45 (81.8%) | 40 (66.7%) | 40 (70.2%) | 0.074 |
| <sup>a</sup> p value for the 4 group comparison (superiority, this does not reflect non-inferiority testing) |  |  |  |  |  |

**Figure 2.**
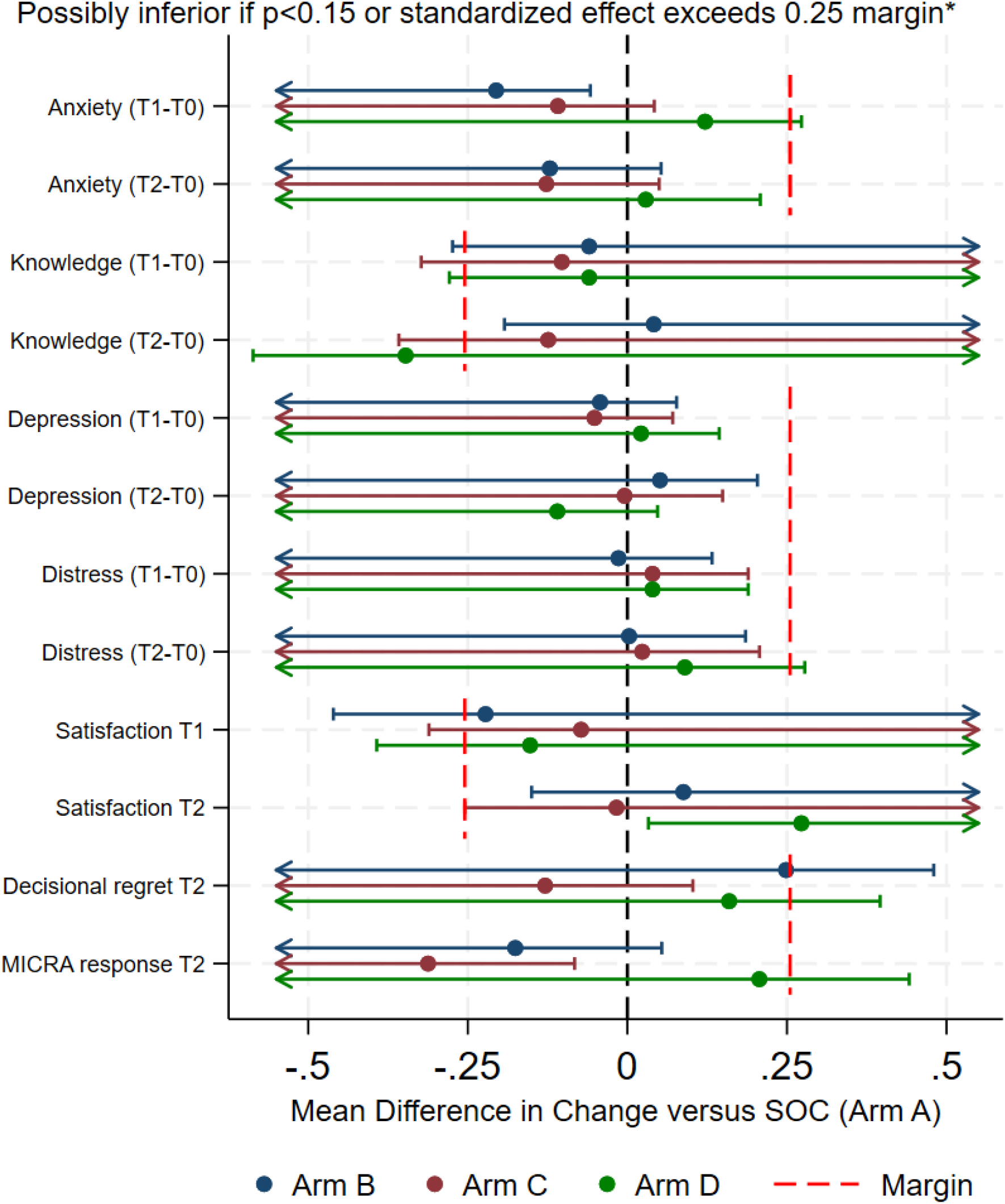
Forest plot depicting prespecified noninferiority margins and confidence intervals by study arm compared to the standard of care arm.

Uptake of visit 1 was significantly lower in arms C and D (p=.01). Uptake of testing was also lower in these arms (p=.07), although uptake of testing did not differ significantly among those who completed a visit 1 (**Table 2**). Participants who did not complete testing were more likely to live in an area of socioeconomic deprivation compared to those who completed testing (SVI 0.4 vs 0.3, p=0.01). There were no other sociodemographic or clinical characteristics associated with testing.

### Requests to speak with a counselor and secondary as-treated analyses

Some participants randomized to a digital visit requested a GC (visit 1-11.1%; visit 2-14.5%)(**Supplemental Table 3**). As-treated non-inferiority analyses were similar to the ITT results for the short-term PROs. Even though some 6-month PROs were non-inferior as discussed earlier, in the as-treated analyses, we met non-inferiority for knowledge in all arms at 6 months. Participants who requested a GC for disclosure in Arm B (GC/Digital) were older (mean age 71.6 years vs 63.3 years, p=0.04), less educated (50% with high school education or less vs 13.5%, p=0.03), more likely to live in a rural area (75% vs 32%, p=0.026), and more likely to receive care at a community site vs academic site (75% vs 16.2%, p=0.003).

Participants in Arm C (Digital/GC) who requested GC for visit 1 had lower health literacy (mean score 4.8 vs 2.0, p=0.002) and were more likely receiving treatment at a community site vs academic (50% vs 12%, p=0.07).Participants who requested a GC for at least one visit in Arm D (Digital/Digital) were older (mean age 72.2 years vs 65.2 years, p=0.063) and had lower baseline genetic knowledge (mean score 6.0 vs 8.7, p=0.019).

### Moderators of patient reported outcomes and testing

For pre-test counseling (visit 1, T0-T1), patients living in poorer areas had greater reductions in anxiety with GCs. Rural patients and those from neighborhoods of lower SVI socioeconomic status had greater increase in knowledge with the digital intervention, while suburban/urban and higher SVI socioeconomic status patients had greater increases with GC counseling. For disclosure visits (T1-T2), men and patients with worse health literacy had greater decreases in anxiety with digital disclosure, while women and those with better health literacy had greater reductions with GC disclosure. Patients from neighborhoods with higher SVI minority status had greater reduction in distress and more positive response with GC disclosure, while those from lower SVI minority status neighborhoods had greater benefits with digital disclosure. There were no significant moderators for uptake of testing.

### Communication of results to relatives and health care providers and impact on clinical care

At 6 months, 45% percent of tested patients intended to share or had shared their results with family members. There were no statistically significant differences in sharing test results by assigned arm or test result. We identified 9 participants with a germline PV (*BRCA1* [1], *BRCA2* [4], prostate cancer: *ATM* [2], *BARD1* [1], *CHEK2* [1]) that could impact treatment. Of these, 5 had disease progression after receiving their results and all had their treatment changed based on genetic testing.

### Genetic counselor time and intervention use

GCs spent the most time on care for participants in Arm A and the least time for participants in Arm D, as shown in **Figure 3**. Specific GC activities and time spent are available in **Supplemental Table 4**. Importantly, even in Arm D (digital/digital), GC spent over 40 minutes coordinating care. The greatest reduction in time was seen in eliminating pretest GC counseling.

**Figure 3.**
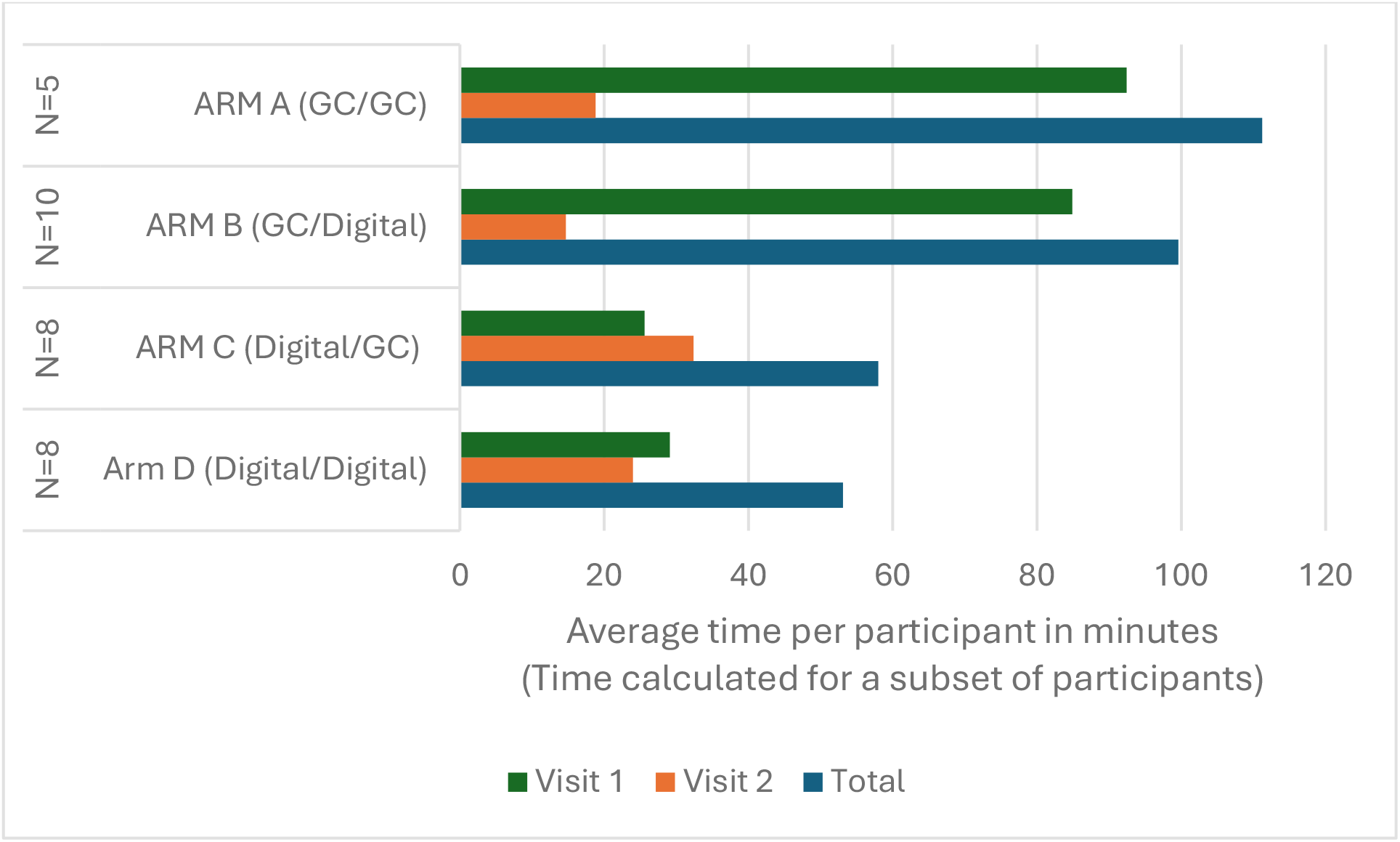
Genetic counselor time per patient by randomized arm and visit.

## DISCUSSION

To our knowledge this is the first randomized non-inferiority study evaluating a digital alternative to GC for pre-test counseling and/or results disclosure in patients with a range of metastatic cancers where germline testing is indicated to inform treatment decisions. We found that offering the eREACH patient-centered digital delivery model with one digital visit and one visit with a GC is non-inferior to two visits with a GC. While the fully digital model may be associated with small differences in short-term knowledge gain, the clinical significance may be small and the fully digital model met non-inferiority for knowledge at 6 months. All digital arms were associated with reductions in GC time, which could increase provider capacity and reduce costs. However, a subset of patients requested counseling with a GC and women and patients with lower health literacy and higher social vulnerability may experience greater short-term benefits from GC visits, highlighting the importance of maintaining this option to provide patient-centered care.

Our findings demonstrating non-inferiority for short-term PROs with both one-visit models (e.g. one GC visit) are similar in some ways to other randomized clinical genetics studies. Two randomized studies among patients with metastatic and/or high risk prostate cancer and one with breast cancer (6.6% metastatic) showed that digital pretest educational tools were non-inferior to counseling with a GC (similar to Arm C in this study).^19–21^ These studies had limited sociodemographic variability and provided genetic testing at no cost to participants. In contrast, our study provides evidence in a real-world population of patients with 4 types of advanced cancers, including patients with less formal education, from rural settings, recruited from community sites and utilized real-world testing practices in which insurance was billed and patients were responsible for any out-of-pocket costs.

Patients with metastatic cancers are often ill at the time of genetic testing and a digital alternative to standard counseling can alleviate the time toxicity associated with 2 GC visits, even when conducted by telehealth. However, genetic education is still important in this population because patients can have different goals - they may want testing to inform treatment options only, or they may want testing to provide information for implication for family members. Our model allowed for different test options to honor these personal preferences and limit uncertainty, whereas most studies include a single option for testing.

While our results were inconclusive regarding whether the fully digital model (with a GC available upon request) was non-inferior for genetic knowledge in the short-term, at 6 months the fully digital model was non-inferior for knowledge in as-treated analyses. Furthermore, the absolute differences between arms were small and in the preferred direction of increased knowledge, and therefore of likely limited clinical significance. Thus, digital pre- and post-test counseling may be acceptable in a carefully selected populations and particularly those where burdens to care may be highest and patients are comfortable with digital alternatives.

Another frequently endorsed delivery model is “Mainstreaming”, wherein the traditional pre-testing counseling session with a GC is replaced with educational materials and a conversation with the oncologist who then orders genetic testing, discloses the results, and offers the patient the option to meet with a GC or refers to a GC when a positive result or VUS is identified. We acknowledge that we cannot directly compare our models to mainstreaming models; But, we can provide direct comparisons to two visits with a GC and there are currently no published studies comparing mainstreaming to two visits with a GC.^22^

Rates of uptake of testing were lower in the digital arms. Notably, uptake of testing was similar across all arms among those who completed visit 1. Additional reminders (which could be automated) or recontacting patients to convert them to a GC visit could address slightly lower uptake of the digital visit 1 in this population. While our testing rates were slightly lower that some other studies^19–21^, many of these covered the cost of testing, which is a common concern for patients and reason for declining testing. Additionally, many of our patients were in rural and community settings, where concerns about cost may be greater. Additional research to understand barriers to testing in patients with metastatic cancer is needed, particularly as all participants with actionable mutations and disease progression had change in systemic treatment based on their test results.

We also identified several subgroups who benefited more or less from digital interventions, including patients from rural areas and neighborhoods with greater social vulnerability, and differences between men and women. Additional research to understand differences in outcomes will be critical for all alternative delivery models if they are incorporated routinely in clinical care. These data and the request for GCs in 13-22% who were assigned to digital visits supports continued access to traditional counseling to address varying patient needs.

We acknowledge some limitations. While this was a prospective, randomized, real-world trial with extensive PROs, 50% enrollment from community settings, and high completion rates, we had slow recruitment and a conditional power analysis was conducted which justified a smaller sample size than originally calculated. Survey response rates were lower for 6-month (T3) outcomes. The proposed model in eREACH requires changes in how GCs practice, especially in the context of post-disclosure visits and broader provider acceptability is unknown. While testing was covered by insurance, development and maintenance of the intervention and GC time was covered by the study and for broader dissemination, these costs will need to be covered by other means including coverage for non-patient provider time requiring new reimbursement models for “virtual care”. However, the provider time savings of digital models demonstrate the potential cost-savings with non-inferior patient outcomes.

In conclusion, the eREACH patient-centered digital delivery intervention with one GC visit is an evidence-based alternative to two visits with a GC for patients with metastatic cancer. The fully digital model may be acceptable for some patients and some subgroups may still benefit from access to a genetic provider to support optimal patient-centered care.

## Data Availability

All data produced in the present study are available upon reasonable request to the authors

## Notes

**Research support:** Primary financial support for this work was supported by the Basser Center for BRCA and a Research Grant from Astrazeneca ESR-19-20239.

### Competing Interest Statement

Primary financial support for this work was supported by the Basser Center for BRCA and a Research Grant from Astrazeneca ESR-19-20239 (ARB).

### Clinical Trial

NCT04353973

### Funding Statement

Primary financial support for this work was supported by the Basser Center for BRCA and a Research Grant from Astrazeneca ESR-19-20239.

### Author Declarations

The Institutional Review Board of the University of Pennsylvania gave ethical approval for this work.

