## Supplemental Tables and Figures for "eREACH: A randomized trial of digital alternatives to genetic counseling for metastatic cancer"

**Table of Contents**

| | **Page Number** |
|---|---|
| Supplemental Table 1: GeM-CheckD: Genomic Medicine Checklist for reporting on Digital tool development, implementation, and evaluation. | 2 |
| Supplemental Table 2: Genetic testing choice by visit 1 completion method. | 6 |
| Supplemental Table 3: Proportion of patients by arm, assigned to a digital visit who requested and completed visit with a genetic counselor | 7 |
| Supplemental Table 4: Average genetic counselor time per participant by arm and activity type | 7 |
| Supplemental Figure 1: Forest plot depicting prespecified noninferiority margin and confidence intervals by study arms compared to usual care group for 6-month outcomes. | 8 |

**Supplemental Table 1:** GeM-CheckD: Genomic Medicine Checklist for reporting on Digital tool development, implementation, and evaluation.

| Domain 1: Digital Tool Development | | | |
|---|---|---|---|
| **Item No.** | **Item Name** | **Description/ sub-Items** | **eREACH** |
| 1.1 | Purpose and Function of the tool | Intent of tool/rationale (e.g., to aid or replace genetic counseling) and function it performs (e.g., help with pre-test or post-test decisions) | To replace traditional pre-test genetic education (visit 1) and/or return of genetic test results (visit 2) with a genetic counselor. Patients still have the option to speak with a counselor. |
| 1.2 | Messaging and Content | Key messages and topics covered by the tool | Visit 1 includes 8 modules which cover purpose of genetic testing and testing options, possible results and implications of results, and testing decision.<br>Visit 2 includes 4 modules which include test results, explanation of result and next steps. |
| 1.3 | Guidelines and/or Theories utilized by the tool | Use of guidelines (e.g., ASCO/ ACMG), frameworks, or models to select content or inform purpose of the tool | Informed by the Tiered-binned model for genetic counseling (Bradbury et al. Gen Med, 2015). |
| 1.4 | Goals/Target of the tool | What is the tool trying to change (e.g., increasing knowledge, reducing decisional conflict, changing behavior) | Provide a digital alternative to reduce the number of scheduled visits for patients and reduce provider time given increasing indications for genetic testing and a limited workforce, while providing equal or better patient outcomes. |
| 1.5 | Intended Users of the tool | Who is the tool for (e.g. patients, providers/ clinicians, general public). May include multiple user types | Patients with advanced or metastatic cancer where germline testing could inform targeted therapy (breast, ovary, pancreas and prostate cancer). |
| 1.6 | Expertise Leveraged | What experts developed and reviewed the Tool | Genetic counselors, medical oncologists, medical ethicist, behavioral scientists, health communication and medical informatics. |
| 1.7 | Tool Characteristics | Name of the tool. Include version used in project/reporting and the latest version available (if applicable) | eREACH1 Digital Intervention |
| | | Delivery mode (e.g. chatbot, website) | Website |

| | | | |
|---|---|---|---|
| | | How information is presented in the tool (e.g. audio/visual, what languages) | Information presented as text, figures, and tables. Optional information includes short videos. Available in English only |
| | | Tool functionalities (e.g. skip logic, interactive, scripted) | Core content is required to be completed by end user. Optional information can be selected to further explore each of the core topics. |
| | | Time for user to complete the tool | ~12-14 mins for visit 2 core (required) content. ~10 mins for visit 2 core content. |
| | | Readability (for written information) | Readability score of $8^{th}$ grade or lower when possible |
| | | Tailoring/Targeting of content (e.g. adaptation within the tool for different individuals, risks, etc) | None |
| | | Use of AI/LLM within the tool – (specify the type of AI/LLM, closed/open/restricted source, algorithm-based, NLP, etc) | None |
| 1.8 | Development and Refinement Process | Involvement of intended users in developing or refining tool (development and post-development) | Tool was refined through user testing with 19 patients and usability testing with 18 patients with advanced cancer |
| | | Methods used to gather feedback (e.g. interviews, user testing) and any measures used for development and refinement | Individual qualitative interviews for user and usability testing. |
| | | Changes/revisions/ adaptations during development and/or during implementation in context | Changes included: simplifying content, changing order and priority of content, increasing representativeness of photos. |
| **Domain 2: Tool Implementation - Processes and Strategies in Context** | | | |
| **Item No.** | **Item** | **Description/ sub-Items** | |
| 2.1 | Implementation Context | How/when/ where tool is accessed by users | Digital intervention is accessed via computer, mobile phone or tablet with a unique user name and password. |
| | | Characteristics of site(s) where tool is deployed | Accessed at home as an alternative to a scheduled visit with a genetic counselor. |
| | | Ethical/ governance / considerations for using the tool by a clinic or health | Individual results are included in the disclosure tool requiring user name, password and confirmation of the user. |

| | | | |
|---|---|---|---|
| | | system (privacy/safety/ security) | |
| 2.2 | Facilitation | Materials (e.g., manuals, SOPs, support tools) | None. |
| | | Ways of selecting or raising awareness among potential users | Not applicable. |
| | | Instructions on tool use (how, when, where, and how often provided) | Instructions on how to use the tool are included at the beginning of the core content. |
| | | Who facilitates use and how | No facilitation required to complete the intervention. |
| | | Engagement and implementation strategies for end users and facilitators (e.g., prompts, incentives, audit/feedback strategies) | Up to 5 reminders sent to prompt use of the tool |
| 2.3 | Implementation or Process Evaluation and Reporting | Methods for evaluating potential impact of context and/or facilitation on outcomes | To be assessed in future research |
| | | Dose (how much exposure or number of times viewed) and fidelity. Quality Control (QC) process or evaluation to determine if tool is used as intended | Designed to require viewing of at least the Tier 1 information. Analysis of differences in outcomes by use are ongoing. |
| | | Costs involved in implementing the Tool | Not assessed at this time |
| | | Opinions or experiences of end users or facilitators during implementation | Not assessed at this time |
| 2.4 | Implementation implications/ information for scale up or broader use of the tool | Evidence suggesting outcomes may be impacted by context or facilitation | Yes: See moderator analyses in this manuscript |
| | | Evidence tool may work differently for different groups of people | Yes: See moderator analyses in this manuscript |
| | | Policy/ practice implications or uses of the Tool broadly | The one-visit arms are non-inferior to two visits with a genetic counselor for all PROs, providing an alternative delivery model. The fully digital model met non-inferiority for all PROs but was inconclusive for knowledge, although the differences are small and likely not clinically significant. Therefore, the fully digital model could be considered for patients comfortable with the digital option. |

| | | | |
|---|---|---|---|
| | | Potential scalability and maintenance (e.g. prerequisite knowledge and skills to use the Tool, funding source to implement the tool, automatic/manual updating of tool and/or guidelines used in tool) | The tool is mobile ready and has been evaluated in real-world and representative patients. It will require maintenance and costs are being determined for broader dissemination. |
| | | Flexibility in implementation (e.g., how easily can the tool adapt to context; are there core elements/processes that can/cannot be adapted) | The tool is not intended to be adapted but changes can be made if needed as clinical care changes. |
| | | Tasks needed for broad dissemination and maintenance | The tool was originally built for only for the research study, but a clinical version is now available upon request. It is also being rebuilt for broader dissemination. |
| | | Remaining gaps in understanding | If outcomes differ among patients with positive and VUS results; further evaluation in non-white patients and how time and costs for virtual or digital care will be covered with clinical implementation. |
| **Domain 3: Digital Tool Outcomes Evaluation** | | | |
| **Item No.** | **Item** | **Description/ sub-Items** | |
| 3.1 | Study/ Evaluation Design & Methods | Details about how the effectiveness/ efficacy of the Tool was tested/determined | Randomized non-inferiority trial as described in this manuscript. |
| | | Regulatory/ IRB: how was the implementation and evaluation approved by the IRB to conduct the study and evaluation | Evaluation study was IRB approved. Informed consent was collected upon invitation to use the tool and provide data for evaluation activities. |
| 3.2 | Study/ Evaluation Population | How participants were selected to use the tool (e.g. eligibility criteria) | Advanced or metastatic breast, ovary, pancreatic or prostate cancer as outlined in this manuscript. |
| | | Incentives provided to participate in the study, complete assessments, or use the tool | No incentive for using the tool. |
| | | Sample size, attrition, participant characteristics | As outlined in the manuscript. |
| 3.3 | Study/ Evaluation Measures & Analyses | Description of outcomes (when measured, what was measured, and what was the | Primary outcomes were knowledge and anxiety. Multiple secondary cognitive, affective and behavioral outcomes and uptake |

| | | | |
|---|---|---|---|
| | | primary outcome of the study) | of pre-test counseling/education and testing. Measured at baseline, after visit 1, after visit 2 and at 6 months. |
| | | Data handling (e.g. missing data protocols, adjustments, consolidation, transformation, etc) | Not applicable |
| | | Types of analyses conducted | Non-inferiority and moderator analyses as described in this manuscript. |
| 3.4 | Results & conclusions of study/ evaluation using the tool | What do results suggest about the tool's ability to achieve its purpose or goals | Non-inferiority supports use of both one-visit models and fully digital model as an alternative to two visits with a genetic counselor in patients with metastatic cancer. |

**Supplemental Table 2**: Genetic testing choice by visit 1 completion method.

| Participant testing choice, n (%) | Visit 1 with GC n=121 | Visit 1 Digital n=77 | |
|---|---|---|---|
| | | | P<.001 |
| No testing | 1 (1) | 1 (1) | |
| Targeted panel[a] | 7 (6) | 21 (27) | |
| Larger panel[b] | 94 (78) | 51 (66) | |
| Custom panel | 19 (16) | 4 (5)[c] | |

[a] *BRCA1, BRCA1* with additional genes tested for patients with prostate cancer: *ATM, BARD1, BRIP1, CDK12, CHEK1, CHEK2, FANCL, PALB2, RAD51C, RAD51D, RAD54L*

[b] Standard Gene Panels for Option 2

– Breast: *ATM, BARD1, BRCA1, BRCA2, BRIP1, CDH1, CHEK2, EPCAM, MLH1, MSH2, MSH6, NBN, PALB2, PMS2, PTEN, RAD51C, RAD51D, STK11, TP53*

– Ovarian: *ATM, BARD1, BRCA1, BRCA2, BRIP1, CHEK2, DICER1, EPCAM, MLH1, MSH2, MSH6, PALB2, PMS2, RAD51C, RAD51D, SMARCA4, STK11, TP53*

– Pancreatic: *ATM, BARD1, BRCA1, BRCA2, BRIP1, CDKN2A, CDK4, CHEK2, EPCAM, MLH1, MSH2, MSH6, PALB2, PMS2, RAD51C, RAD51D, STK11, TP53, VHL*

– Prostate: *ATM, BARD1, BRCA1, BRCA2, BRIP1, CHEK2, EPCAM, HOXB13, MLH1, MSH2, MSH6, NBN, PALB2, PMS2, RAD51C, RAD51D, TP53*

[c] Percentages may not add to 100 due to rounding

**Supplemental Table 3:** Proportion of patients by arm, assigned to a digital visit who requested and completed visit with a genetic counselor

| | Arm B (GC/Digital) | Arm C (Digital/GC) | Arm D (Digital/Digital) |
|---|---|---|---|
| Visit 1 (pre-test) | N/A | 6/46 (13.0%) | 7/47 (14.9%)[a] |
| Visit 2 (disclosure) | 8/45 (17.8%) | N/A | 9/40 (22.5%) |
| [a]Among patients in Arm D, 1 requested a GC for visit 1 only, 3 requested a GC for visit 2 only, and 6 requested a GC for both visits. | | | |

**Supplemental Table 4** : Average genetic counselor time per participant by arm and activity type

| **Visit 1 Activity** Average time per participant in mins (range) | Arm A (GC/GC) N=5 | Arm B (GC/Digital) N=10 | Arm C (Digital/GC) N=8 | Arm D (Digital/Digital) N=8 | Average time per participant by activity |
|---|---|---|---|---|---|
| Chart review | 9 (0-15) | 13.2 (7-20) | 5 (3-10) | 7.9 (3-14) | 9 (0-20) |
| Session time | 54 (37-84) | 48.2 (26-90) | NA | NA | 50.1 (26-90) |
| Communication with patient/provider after session | 1 (0-5) | 2.1 (0-10) | 3.6 (0-15) | 1.5 (0-12) | 2.2 (0-15) |
| Facilitating testing | 11.6 (8-15) | 10.6 (4-20) | 9 (5-15) | 10.9 (6-20) | 10.4 (4-20) |
| Insurance Navigation | 4.4 (0-12) | 0.5 (0-5) | 0.8 (0-6) | 0.2 (0-2) | 1.2 (0-12) |
| Documentation | 10 (0-15) | 9.8 (0-20) | 5.4 (4-10) | 7.1 (0-12) | 8.2 (0-20) |
| Other | 2.4 (0-12) | 0.5 (0-5) | 1.9 (0-15) | 0.6 (0-5) | 1.2 (0-15) |
| V1 total average time per participant | 92.4 (67-151) | 84.9 (46-145) | 25.6 (14-40) | 29.1 (17-40) | |
| **Visit 2 Activity** | Arm A (GC/GC) N=4* | Arm B (GC/Digital) N=10 | Arm C (Digital/GC) N=8 | Arm D (Digital/Digital) N=8 | Average time per participant by activity |
| Approval of web result | NA | 7.3 (4-10) | NA | 4.7 (3-7) | 6.4 (3-10) |
| Chart review | 1.2 (0-5) | 1.4 (4-5) | 1.1 (0-4) | 2 (0-5) | 1.5 (0-5) |
| Session time | 7.2 (4-9) | 5 (5) | 10.8 (6-20) | NA | 9.2 (4-20) |
| Communication with patient/provider after session | 2.5 (0-10) | 0.6 (0-2) | 0.6 (0-2) | 0.5 (0-2) | 0.5 (0-2) |
| Documentation | 11.2 (5-15) | 9.7 (7-13) | 14.7 (10-23) | 11.9 (5-21) | 11.7 (5-23) |
| Other | 0 | 0 | 4.9 (0-25) | 1.5 (0-12) | 1.7 (0-25) |

| | | | | |
|---|---|---|---|---|
| V2 total average time per participant | 18.8 (13-33) | 14.7 (11-18) | 32.4 (18-59( | 24 (9-33) |
| Combined V1 and V2 GC time | 111.2 (170-180) | 99.6 (63-160) | 58 (40-99) | 53.1 (31-73) |
| *One participant in Arm A did not have testing<br>NA=not applicable | | | | |

**Supplemental Figure 1**: Forest plot depicting prespecified noninferiority margin and confidence intervals by study arms compared to usual care group for 6-month outcomes.

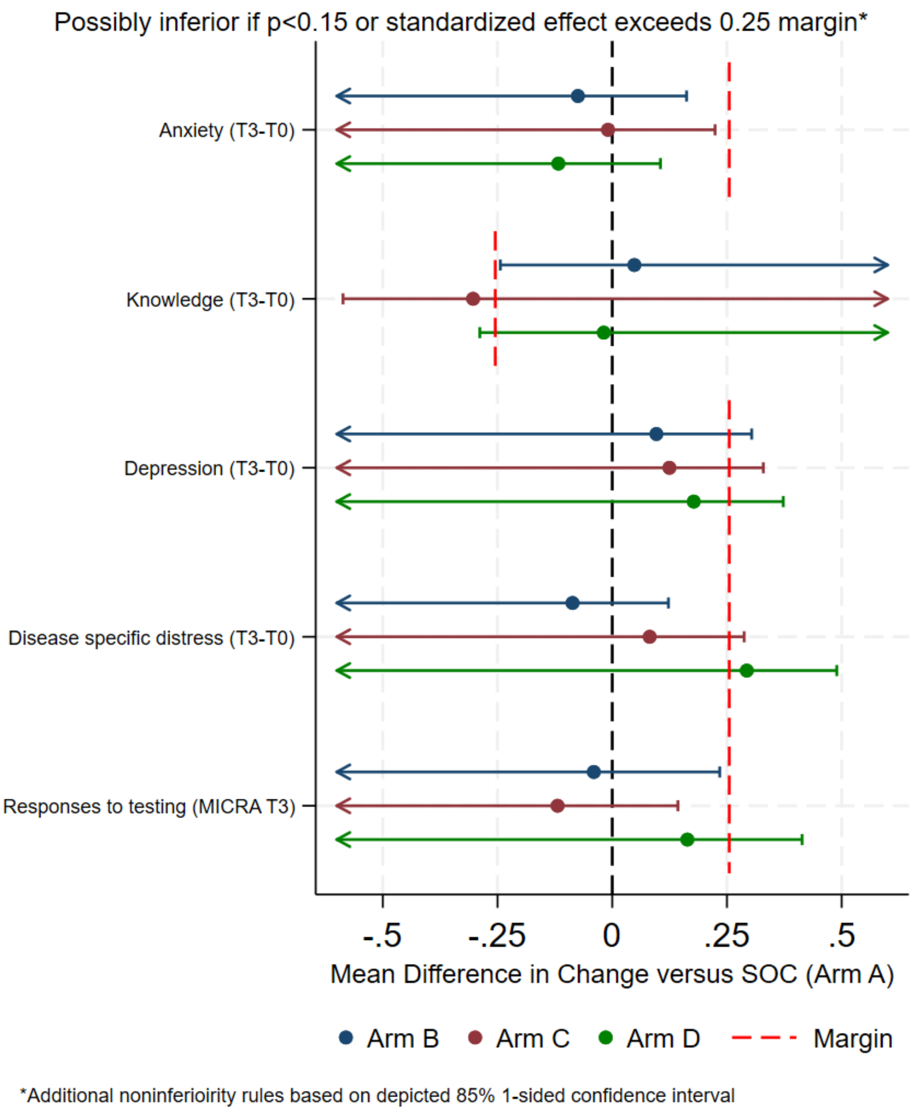
